# Prediction and Prevention of Ventilation Impairments During Bronchoscopy

**DOI:** 10.1101/2025.05.12.25327413

**Authors:** Ben Fabry, Navid Bonakdar, Christian Kuster, Johannes Bartl, Frederick Krischke, Roland Francis

## Abstract

Bronchoscopy in mechanically ventilated patients is performed by passing a bronchoscope through the endotracheal tube (ETT), which substantially increases airflow resistance and may compromise ventilation. Here, we quantify the nonlinear, flow-dependent resistance of ETTs with and without a bronchoscope by analyzing pressure-flow relationships across multiple tube-bronchoscope configurations. We find that with bronchoscope insertion, tube resistance increases with the inverse fifth power of the effective tube diameter, defined as the diameter of a circular tube with the same cross-sectional area as the remaining lumen. Using an intensive care ventilator in combination with an active lung simulator, we demonstrate that the increased resistance during bronchoscopy causes dynamic hyperinflation and intrinsic positive end-expiratory pressure (PEEP) build-up in volume-controlled modes, and reduced tidal volumes in pressure-controlled modes. Numerical simulations using a simple scaling law relating resistance to effective tube diameter accurately reproduce the observed impairments. This demonstrates that the impact of tube narrowing during bronchoscopy can be reliably predicted from ventilator settings and patient respiratory mechanics. We present a predictive model that allows clinicians to anticipate and manage ventilation impairments, supporting evidence-based selection of endotracheal tubes and bronchoscopes. In addition, we provide proof of principle that combining pressure-controlled ventilation with automatic tube compensation can fully prevent these impairments, pointing to a technically feasible solution to an underrecognized clinical problem.

## Introduction

Mechanical ventilation remains a cornerstone in the management of critically ill patients, providing life-sustaining respiratory support in more than 30% of all intensive care unit admissions [1, 2]. In over 90% of these patients, ventilation is delivered invasively via endotracheal intubation to ensure reliable access to the airways [1]. However, intubation is associated with several adverse effects, including vocal cord injury, damage to the tracheal mucosa, and impaired mucociliary clearance, often necessitating frequent airway suctioning. In addition, bacterial biofilms readily form on the inner surface of the tube, creating a conduit for bacterial colonization of the lungs [3].

Beyond these well-known complications, the endotracheal tube introduces substantial and nonlinear airflow resistance [4], which imposes a significant mechanical burden on respiratory function [5]. The resulting flow limitations can resemble the airflow pattern seen in patients with obstructive airway diseases such as asthma [5, 6]. As a consequence, depending on the tube’s internal diameter, there is a risk of dynamic hyperinflation and intrinsic positive end-expiratory pressure (iPEEP) build-up during controlled mechanical ventilation [6]. In addition, elevated resistance contributes to increased work of breathing and patient-ventilator asynchrony during ventilation modes that support spontaneous breathing efforts [7, 8].

Airflow limitations are further amplified when a bronchoscope is inserted through the endotracheal tube, which substantially decreases the available lumen and increases its resistance. Flexible bronchoscopy is a common diagnostic and therapeutic procedure in mechanically ventilated intensive care patients. More than 10% of them undergo at least one bronchoscopic intervention during their stay, with higher frequencies in patients requiring prolonged ventilation or experiencing respiratory failure [9]. Common indications include evaluation of persistent infiltrates, removal of mucus plugs, and bronchoalveolar lavage [10]. Bronchoscopy is typically performed through the endotracheal tube via a sealed swivel adapter, allowing it to proceed without disconnecting the ventilator.

Alterations in airflow dynamics during bronchoscopy can be substantial, especially for combinations of small endotracheal tubes and large bronchoscopes. In such cases, the intrinsic PEEP build-up in volume-controlled modes and the reduction of tidal volume in pressure-controlled modes can reach critical levels [11, 12]. However, these effects are also dependent on ventilator settings and the patient’s respiratory system elastance [11, 12]. Consequently, continuous bedside estimation of iPEEP and tidal volume is mandatory during bronchoscopy, but remains technically challenging and prone to error. Compensation strategies that dynamically adjust airway pressure to account for increased tube resistance, such as Automatic Tube Compensation (ATC), may offer a potential solution, although they have not yet been evaluated during bronchoscopic procedures.

The aim of the current study is to characterize the nonlinear flow-dependent resistance of different tube-bronchoscope combinations and to develop a quantitative framework for accurately predicting iPEEP and tidal volume based on ventilator settings and patient respiratory mechanics. In addition, we evaluate whether Automatic Tube Compensation can prevent ventilation impairments during bronchoscopy.

## Methods

For measuring the resistance of original-length endotracheal tubes (Rüsch Super Safety Clear, Teleflex, Ireland; inner diameter 6 to 9 mm in 0.5 mm increments), we replicated the setup described in [4] (Fig. 1). In brief, we inserted the tip of the endotracheal tube and cuffed it airtight in an artificial trachea (Plexiglas tube) with an inner diameter of 2.1 cm. Tracheal pressure was measured through 12 small, equally spaced radial holes positioned around the circumference of the artificial trachea, located 60 mm distal to the tip of the endotracheal tube and 50 mm proximal to the end of the artificial trachea (Fig. 1). Similarly, airway pressure was measured through 8 small, equally spaced radial holes in a tube placed between the swivel connector and the flow meter.

**Fig. 1:**
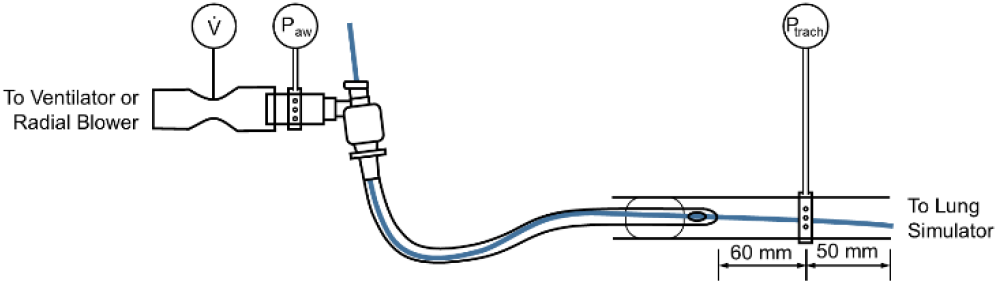
Setup for measuring the influence of a bronchoscope (blue) on tube resistance, flow, and pressure pattern during mechanical ventilation. The tube is inserted in an “artificial trachea” with a diameter of 2.1 cm. The bronchoscope is inserted through the tube beyond the measurement position of the tracheal pressure, P_trach_.

Airway and tracheal pressures were measured with piezo-resistive differential pressure sensors (HCS-series Honeywell (USA) sensors with SPI serial interface, ±80 mbar range). Gas flow was measured with a factory-calibrated thermal mass flow sensor (SFM3300, Sensirion, Switzerland). Flow and pressure signals were sampled at 250 Hz and digitally low-pass filtered using a 4th-order Bessel filter with a cutoff frequency of 25 Hz.

The artificial trachea was connected to a custom-built active lung simulator consisting of a 2.8 L syringe with a piston driven by a stepper motor via a spindle-nut mechanism. The simulator applies the equation of motion of the respiratory system at an update rate of 250 Hz. Respiratory system compliance was set to 50 ml/mbar and airway resistance to 2 mbar/(L/s). Muscular pressure was set to zero to emulate the absence of spontaneous breathing efforts.

For measuring the tube resistance, the lung simulator was removed, and a radial blower (U65HN-024KS-6, Micronel, Switzerland) was connected to the flow meter. The motor speed of the radial blower was slowly ramped up and down to generate a maximum pressure (at zero flow) of +80 mbar (at the blower outlet), or −80 mbar (at the blower inlet). Eq. 1 was fitted to the measured pressure-flow relationship across the tube using least-squares optimization implemented in the SciPy library of Python.

For exploring the pressure and flow patterns during pressure- and volume-controlled mechanical ventilation without automatic tube compensation, we used an intensive care ventilator (EVITA V600, Dräger, Germany) in combination with the lung simulator. Ventilator settings during volume-controlled ventilation were: tidal volume *V*_T_ = 500 ml, positive end-expiratory pressure PEEP = 0, inspiratory time T_in_= 1.8 s without end-inspiratory pause, expiratory time T_ex_ = 2.2 s. Parameters during pressure-controlled ventilation were: Pressure support = 10 mbar above PEEP, PEEP = 0, T_in_= 1.8 s, T_ex_ = 2.2 s, pressure ramp time = 0.

To deliver pressure-controlled mechanical ventilation with automatic tube compensation (ATC), we used a prototype ventilator based on the system described in [8], with the following modifications: positive and negative pressures were generated by two radial blowers (U65HN-024KS-6, Micronel, Switzerland) connected to the inspiratory and expiratory limbs of a Y-piece, respectively. The patient limb of the Y-piece was then connected to the flow meter (Fig. 1). Pressure was controlled by a 3-way valve integrated directly into the Y-piece, as described in [13]. Ventilator settings were as follows: PEEP = 0 mbar, T_in_= 1.8 s, T_ex_ = 2.2 s. Over the course of the inspiratory phase, the target tracheal pressure was linearly ramped up from 0 mbar to 10 mbar. Over the course of the expiratory phase, the target tracheal pressure was linearly ramped down from 10 mbar to 0 mbar.

We performed measurements with single-use bronchoscopes (aScope 4 Broncho, Ambu, Ballerup, Denmark) in three sizes - small, medium, and large - with nominal outer diameters of 4.3 mm, 5.3 mm, and 6.3 mm, respectively. These nominal values refer to the maximum diameter measured at the distal tip of the bronchoscope. However, since the tip extends well beyond the endotracheal tube during use, its diameter does not significantly affect the effective tube resistance. More relevant is the average outer diameter of the bronchoscope shaft along the segment residing within the tube, which is smaller: 3.8 mm, 5.0 mm, and 5.9 mm for the small, medium, and large bronchoscopes, respectively. Accordingly, all calculations in this study are based on these measured shaft diameters rather than the nominal tip diameters. Bronchoscopes were introduced into the endotracheal tube via the swivel connector cap through a perforated silicone membrane seal. To prevent air leakage during use of the smallest bronchoscope, the membrane opening was additionally sealed with adhesive putty (Patafix, Bolton Adhesives).

Tidal volume was computed by numerically integrating the measured flow during inspiration. Intrinsic PEEP was estimated breath-by-breath from the end-expiratory tracheal pressure, which is a close proxy for the alveolar pressure under the conditions of a small end-expiratory gas flow and a small airway resistance of 2 mbar/(L/s).

Numerical simulations of flow and pressure patterns during pressure-controlled and volume-controlled ventilation were performed using a custom Python script, freely available under the MIT license at https://github.com/fabrylab/Bronchoscopy. This repository also includes a browser-based version of the simulation program, implemented in HTML and JavaScript, which can be downloaded and run locally in a web browser, or accessed directly via https://fabrylab.github.io/Bronchoscopy/.

## Results

### Resistance of the endotracheal tube

The pressure difference across the endotracheal tube Δp_ETT_, measured as the difference between the airway pressure p_aw_ and the tracheal pressure p_trach_ (Fig. 1) shows a strong non-linear increase with gas flow, 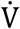 (Fig. 2). This non-linear pressure versus flow relationship is well captured by the Rohrer equation with two free parameters, k1 and k2 [4, 14, 15] (Fig. 1):

**Fig. 2:**
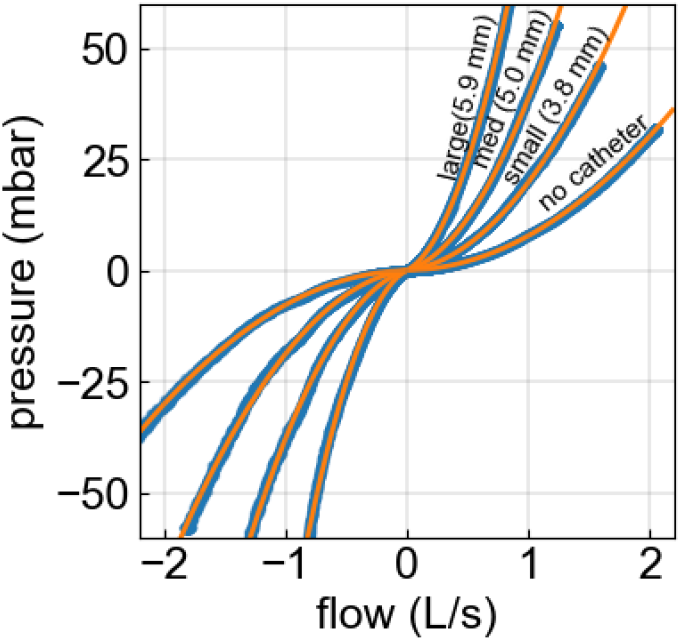
Pressure-flow relationship of an 8 mm inner diameter endotracheal tube before and after inserting bronchoscopes with different outer diameters. Blue dots are measured values; orange lines represent the fit of Eq. 1 to the data.

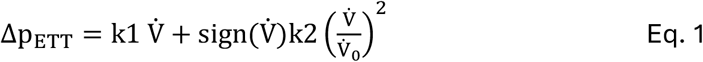

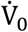 is a reference flow, which we set to 1 L/s so that k1 + k2 corresponds to the total pressure drop at a flow of 1 L/s. k1 describes the fraction of the total pressure drop across the tube that grows linearly with flow, and k2 describes the fraction that grows quadratically with flow. k1 is considered to be the same in inspiration and expiration, while k2 can be different in inspiration and expiration due to the compression-expansion asymmetry of the flow at the transition between the tip of the endotracheal tube and the trachea [4]. Using separate k2-parameters in inspiration and expiration significantly improves the fit of Eq. 1 to the data for a given tube-bronchoscope combination. When comparing different tube-bronchoscope sizes and combinations, however, k2 does not systematically differ between inspiration and expiration (Table S1). Values for k1 and k2 for all combinations of endotracheal tube diameters and bronchoscope diameters are given in Table S1, with k2 being listed separately for inspiration and expiration.

In the following, when we refer to numerical values of resistance R, we specifically mean the secant resistance of the tube at a flow of 1 L/s (the pressure drop at 1 L/s divided by the flow), and hence we report the numerical values of R = k1+k2, averaged for inspiration and expiration, in units of mbar/(L/s). We find that the resistance of endotracheal tubes without inserted bronchoscope increases with the inner tube diameter D according to a power-law: R ~ D^−3.6^ (Fig. 3, gray points and line).

**Fig. 3:**
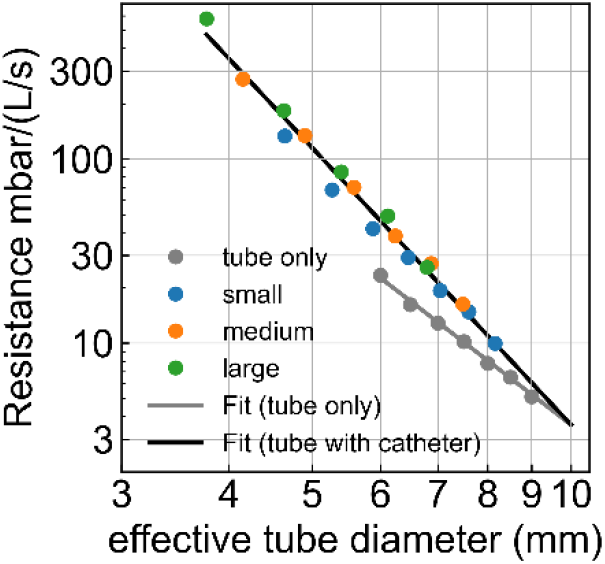
Tube resistance (pressure drop across the tube measured at a flow of 1 L/s, as taken from the Rohrer parameters k1+k2 (Eq. 1) averaged for inspiration and expiration), as a function of the effective tube diameter D_eff_. Points indicate measured values, the gray line indicates the fit R = 3.6 (D_eff_/D_0_)^−3.6^, and the black line the fit R = 3.6 (D^eff^/D_0_)^−5^, with reference tube diameter D_0_ set to 10 mm.

### Tube resistance during bronchoscopy

Inserting a bronchoscope into an endotracheal tube dramatically increases the resistance (Fig. 2), as indicated by an increase both of the linear and the quadratic resistance parameters k1 and k2 (Fig. 3, Table S1). Instead of an inverse power-law relationship with exponent −3.6, however, the resistance increases with the effective diameter D_eff_ according to R ~ D_eff_^−5^ (Fig. 3), whereby D_eff_ is the diameter of a circular tube with the same cross section as the tube-bronchoscope combination:

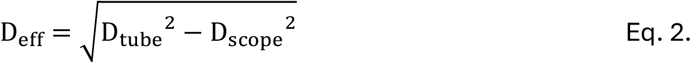

**Table 1.** Effective inner diameters (mm) of endotracheal tubes with and without bronchoscope.

| ETT ID<br>(mm) | Bronchoscope sizes |  |  |
| --- | --- | --- | --- |
|  | Small<br>(3.8 mm) | Medium<br>(5.0 mm) | Large<br>(5.9 mm) |
| 9 | 8.2 | 7.5 | 6.8 |
| 8.5 | 7.6 | 6.9 | 6.1 |
| 8 | 7.0 | 6.3 | 5.4 |
| 7.5 | 6.5 | 5.7 | 4.6 |
| 7 | 5.9 | 5.0 | 3.8 |
| 6.5 | 5.3 | 4.3 | - |
| 6 | 4.6 | - | - |

ETT ID = endotracheal tube internal diameter. Bronchoscope sizes refer to their outer measured diameter, not the nominal diameter provided by the manufacturer.

### Effects of tube resistance on dynamic hyperinflation during volume-controlled ventilation

A numerical simulation of the flow conditions during volume-controlled mechanical ventilation predicts that the intrinsic PEEP increases sharply as the effective tube diameter falls below 5 mm, even for a rather moderate situation with relatively low tidal volume (500 ml), low respiratory rate (15/min), and long (2.2 s) expiration time (Fig. 4a). The Python-script for performing the numerical simulation is provided as supplementary information. We also provide a web-based implementation of the simulation program, written in Java-script, at https://fabrylab.github.io/Bronchoscopy/.

**Fig. 4:**
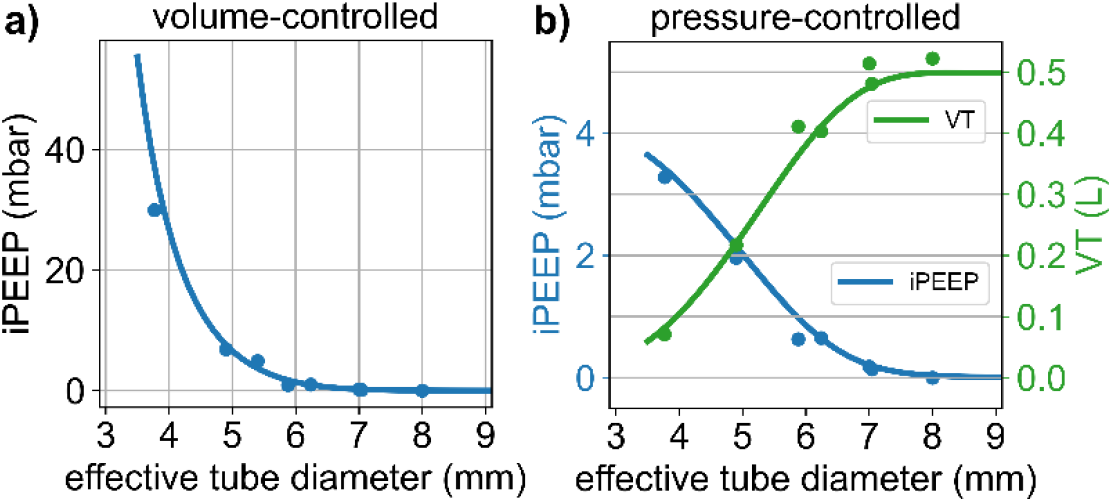
**a)** Intrinsic PEEP build-up during volume-controlled mechanical ventilation, for V_T_ = 500 ml, T_in_= 1.8 s and T_ex_ = 2.2 s, as a function of the effective tube diameter. **b)** Intrinsic PEEP build-up (blue) and tidal volumes (green) during pressure-controlled mechanical ventilation for a pressure support of 10 mbar, T_in_= 1.8 s and T_ex_ = 2.2 s, as a function of the effective tube diameter. In both graphs, the lines show the results from a numerical simulation based on the resistances of tubes with inserted bronchoscope taken from the fit shown in Fig. 3 (black). Dots are measured values obtained with an EVITA V600 ventilator and lung simulator (C = 50 ml/mbar, R_aw_= 2 mbar/(L/s) for different tube-bronchoscope combinations.

The results of the numerical simulations closely agree with measurements of intrinsic PEEP obtained with an EVITA V600 ventilator and a lung simulator for different combinations of endotracheal tube and bronchoscope diameters (Fig. 4a). Therefore, the effects of an increased tube resistance during bronchoscopy can be accurately predicted in advance, based on the ventilator settings, the patient’s respiratory mechanics, and the effective tube diameter. The pressure and flow traces (Fig. 5 top row) illustrate the dramatic airway pressure increase during inspiration and the progressive worsening of expiratory flow limitation with increasing bronchoscope diameter.

**Fig. 5:**
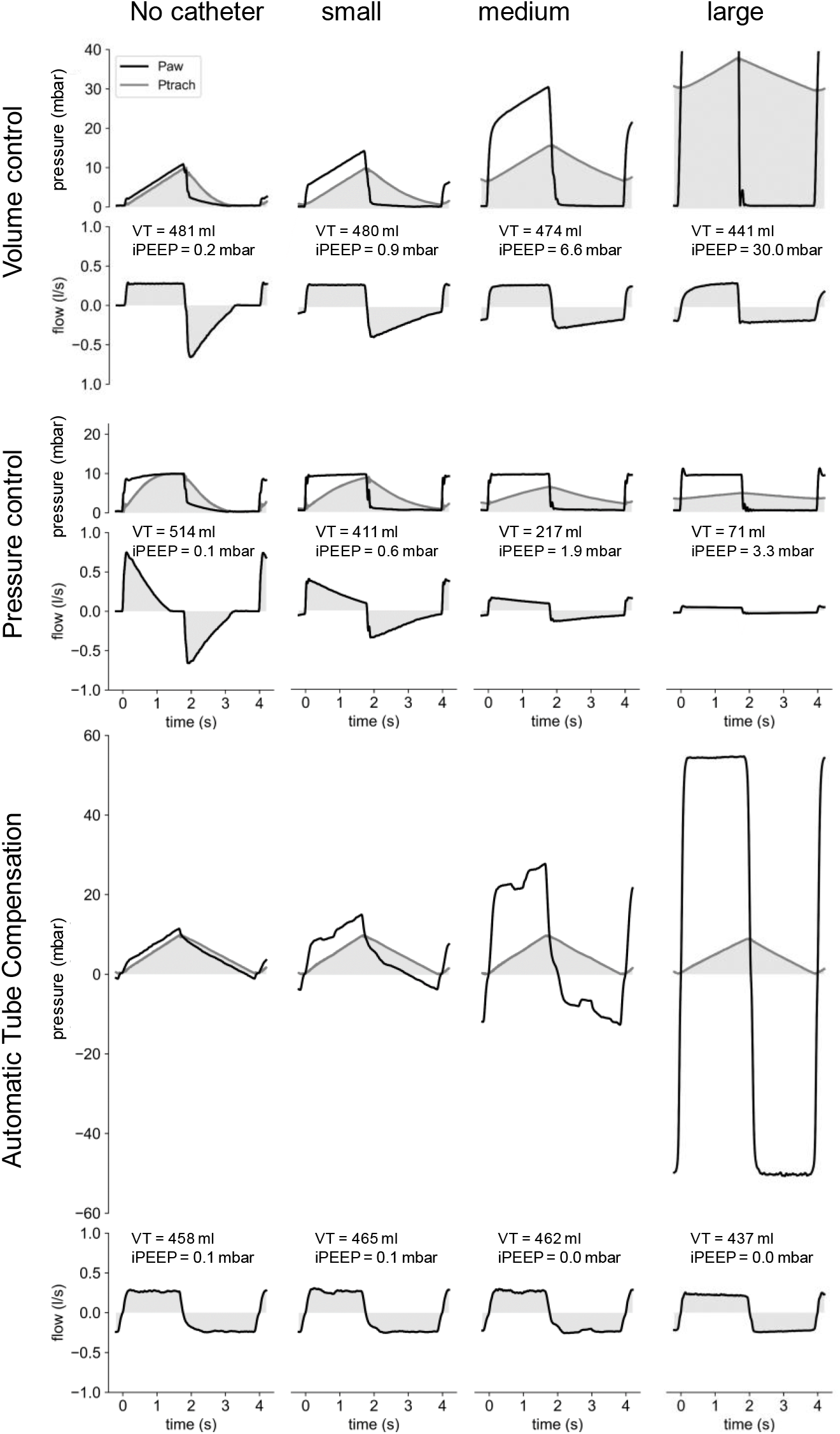
Airway pressure, tracheal pressure, and flow curves during volume-controlled ventilation (top), pressure-controlled ventilation without ATC (middle), and pressure-controlled ventilation with ATC (bottom) through a 7.0 mm tube without or with a small, medium or large bronchoscope. Pressure and flow is delivered by a commercial intensive care ventilator or an ATC prototype ventilator, with settings as described in Methods. The tube and artificial trachea is connected to an active lung simulator. Note that airway pressure exceeded the sensor’s 80 mbar range in VC mode for the large bronchoscope.

As the total resistance increases, the airway pressure can reach very high values (Fig. 5). Due to the compressibility of the ventilator tubing and the gas it contains, a fraction of the delivered flow is needed for pressurizing the system during the initial phase of inspiration (Fig. 5), as can be seen by the more gradual onset of inspiratory flow as bronchoscope sizes increase. Consequently, the tidal volume decreases from 496 ml for an unobstructed 7 mm tube to 441 ml when a large (5.9 mm) bronchoscope is inserted. This slight decline in tidal volume is not taken into consideration in our numerical simulation.

### Effects of tube resistance on tidal volume during pressure-controlled ventilation

The increase of the tube resistance during bronchoscopy not only causes flow limitation during expiration but also limits the inspiratory flow in pressure-controlled modes of ventilation (Fig. 5), leading to progressively decreasing tidal volumes for smaller effective tube diameters. Because of the decreasing tidal volumes, the intrinsic PEEP build-up is smaller in pressure-controlled compared to volume-controlled modes, and will not exceed the driving pressure times the ratio of T_in_/(T_in_+T_ex_), assuming the inspiratory and expiratory valve resistances are not markedly different.

To estimate the intrinsic PEEP and tidal volume changes as a function of the effective tube diameter, we again perform a numerical simulation. The settings for inspiratory and expiratory time, valve resistance and respiratory system mechanics are the same as used above for simulating volume-controlled ventilation, except that we now supply a pressure support of 10 mbar with every breath. We find that for effective tube diameters above 7 mm, the resulting tidal volumes are 500 ml, without noticeable intrinsic PEEP build-up. Below 7 mm, tidal volume decreases, and intrinsic PEEP increases with smaller effective tube diameters (Fig. 4b). The results from the numerical simulations again closely agree with measurements of intrinsic PEEP and tidal volumes obtained with a ventilator and a lung simulator for different combinations of endotracheal tube and bronchoscope diameters (Fig. 4b). The flow traces (Fig. 5) illustrate the progressive worsening of both inspiratory and expiratory flow limitation with increasing bronchoscope diameter.

### Effects of the respiratory system compliance on tidal volume and iPEEP

The increased resistance R of the tube-bronchoscope combination increases the time constant τ of the expiratory flow according to τ = R*C*, with R being the resistance and *C* the compliance of the respiratory system. Because of the non-linearity of the tube resistance, the expiratory flow is not strictly exponential, and the relationship holds only approximately. Nonetheless, even a moderate decrease of the tube cross section will lead to a noticeable lengthening of the time needed to exhale a previously inhaled tidal volume *V*_T_.

When τ approaches the expiration time T_ex_, the expiration will be incomplete, leading to dynamic hyperinflation of the lungs when a constant tidal volume is enforced under volume-controlled ventilation. On the one hand, an increased compliance and hence increased time constant exacerbates iPEEP. On the other hand, a higher compliance reduces the peak alveolar pressure according to Palv,peak = PEEP+*V*_T_/*C*, which tends to lower the end-inspiratory alveolar pressure and thereby reduces iPEEP. These two opposing effects nearly cancel out, such that the build-up of iPEEP at small effective tube diameters is largely independent of the patient’s compliance under volume-controlled ventilation (Fig. SI 1a).

The effects of compliance changes are more pronounced under pressure-controlled ventilation. To maintain a desired tidal volume target *V*_T,target_, the inspiratory pressure support (PS) above PEEP is adjusted to lower values in patients with higher compliance, according to PS = *V*_T,target_ / *C*. This also limits the intrinsic PEEP, which cannot exceed the level of pressure support. However, with increasing compliance, the time constant for filling and emptying the lungs is increased, which tends to increase the intrinsic PEEP and decrease the tidal volume. The latter effect dominates the response, and the effective diameter at which the tidal volume starts to decline shifts to higher values as the compliance increases (Fig. SI 1b).

### Effects of respiratory rate on tidal volume and intrinsic PEEP

Increasing the respiratory rate is associate with increased minute ventilation and decreased time for inspiration and expiration. Not surprisingly, this leads to higher intrinsic PEEP values under volume-controlled modes, and lower tidal volumes under pressure-controlled modes (Fig. SI 2).

### Automatic compensation of tube resistance

Under the mode Automatic Tube Compensation (ATC), the ventilator automatically increases or decreases the airway pressure to whatever value is needed for delivering a target tracheal pressure. This way, the flow-limiting effect of the tube resistance can be prevented. During expiration, it may be necessary to lower the airway pressure below PEEP, or even below atmospheric pressure, to maintain a positive tracheal pressure. Since commercial ventilators, to the best of our knowledge, are not equipped to supply sub-atmospheric pressure, we use a custom-built ventilator for testing if ATC is able to compensate for the increased tube resistance during bronchoscopy, and to maintain a desired tidal volume without intrinsic PEEP build-up.

When the ventilatory support under the mode ATC is delivered in the form of a linear tracheal target pressure ramp (Fig. 5), we find that the inspiratory airway pressure generated by the ATC ventilator resembles the airway pressure during volume-controlled ventilation (Fig. 5). Hence, regardless of the tube resistance, the tidal volume remains approximately constant, except when the airway pressure required to compensate for the tube resistance exceeds the maximum pressure that the ventilator can deliver (Fig. 5).

During expiration, the airway pressure required to compensate for the added tube resistance during bronchoscopy reaches sub-atmospheric levels, while the tracheal pressure remains equal or above PEEP. Lowering airway pressure during expiration prevents any intrinsic PEEP build-up. Taken together, our measurements confirm that the mode Automatic Tube Compensation can effectively compensate for the added resistance during bronchoscopy and prevent both hypoventilation and intrinsic PEEP build-up.

## Discussion

Bronchoscopic procedures in mechanically ventilated patients typically involve the insertion of a bronchoscope through the endotracheal tube (ETT), which significantly increases airflow resistance and may compromise effective ventilation. The current study quantifies the flow resistance of ETTs both with and without bronchoscope, providing insight into how the resulting changes of tube resistance impact ventilatory dynamics. These findings are critical for understanding how the partial tube obstruction during bronchoscopy affects mechanical ventilation, particularly in terms of tidal volume and intrinsic positive end-expiratory pressure (iPEEP), under various ventilator settings. Below, we discuss the key factors influencing ventilation during bronchoscopy.

The results of the current study indicate that the effective diameter of the tube-bronchoscope combination is the sole parameter of practical relevance in defining flow resistance during bronchoscopy. The effective diameter is defined as the diameter of a circular tube that has an equivalent cross-sectional area as the remaining lumen of the tube-bronchoscope system. The effective tube diameter and the resulting tube resistance critically determine the airflow and pressure patterns during mechanical ventilation. Importantly, different combinations of endotracheal tubes and bronchoscopes yielding the same effective diameter will produce the same flow and pressure profiles when ventilator settings are held constant. As such, this effective diameter provides a practical metric for predicting ventilatory outcomes, including intrinsic PEEP and tidal volume, for a given ventilator setting.

Our study further demonstrates that the resistance of an unobstructed endotracheal tube scales with the inner tube diameter D according to a power-law relationship with exponent −3.6. This finding is somewhat unexpected, as the gas flow in endotracheal tubes is predominantly turbulent [4], where a *D*^−5^ scaling would be expected [15]. The observed *D*^−3.6^ scaling for unobstructed tubes is likely explained by the fact that the endotracheal tube is only one component of the full flow path, which also includes the transition into a 2.1 cm-diameter tracheal segment and a 90° swivel connector. These additional elements introduce significant pressure losses due to flow separation and curvature, which are less sensitive to tube diameter [4] and therefore reduce the apparent scaling exponent.

By contrast, if a bronchoscope is inserted, the resistance becomes dominated by frictional losses in the tube. Experimentally, we find that the tube resistance scales as expected by theory with the effective tube diameter according to D_eff_^−-5^. This stronger scaling exponent compared to an unobstructed tube likely results from the highly non-circular flow geometry introduced by the bronchoscope and from additional wall shear stresses along the bronchoscope surface, thereby enhancing frictional losses beyond those of a smooth-walled circular tube with equal cross-sectional area.

The pressure drop across a tube - with or without a bronchoscope - increases non-linearly with flow rate. This relationship is well captured by the Rohrer equation [14], which consists of a component of the pressure drop that increases linearly with flow rate, k1, and a component that increases quadratically with flow rate, k2 [14, 15]. The coefficient k2 for any given measurement also depends on the flow direction, hence, k2 can differ between inspiration and expiration, however, we find no systematic trend for different tube-bronchoscope combinations. Moreover, k2 tends to increase as k1 increases, hence k1 and k2 are co-variant. On average, we find that k2 = 4 k1. This reduces the degrees of freedom and allows us to approximately describe the pressure drop across the tube with a single free parameter - the effective tube diameter - according to

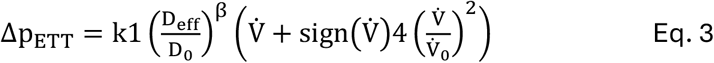

with k1 = 0.72 mbar, and exponent β = −3.6 for an unobstructed tube (without bronchoscope), and β = −5 for a tube with inserted bronchoscope. D_0_ is a reference diameter set to 10 mm. This relationship holds for any tube-bronchoscope combination, for tube diameters between 6.0 - 9.0 mm, and bronchoscope diameters between 3.8 - 5.9 mm. The mean absolute percentage error of Eq. 3 for describing the resistance of unobstructed tubes is 2.4%, and 10.6% for describing the resistance of tubes with inserted bronchoscope.

This universal relationship with only one free parameter, D_eff_, allows us to approximately predict the effects of different endotracheal tube-bronchoscope combinations on the flow and pressure profile during volume-controlled or pressure controlled ventilation, and to compute the resulting intrinsic PEEP and tidal volumes depending on the ventilator settings and the patient’s respiratory system mechanics (Fig. 4). Because of the non-linearity of the pressure-flow relationship across the tube, this computation requires a numerical simulation. We have confirmed that the predictions from the numerical simulation closely agree with measurements obtained with an intensive care ventilator and a lung simulator, for different tube-bronchoscope combinations.

Our study identifies the effective diameter as the crucial parameter that determines intrinsic PEEP build-up in volume-controlled modes, and diminished tidal volumes in pressure-controlled modes. Conclusions from earlier studies that the endotracheal tube diameter should be at least 2 mm larger than the diameter of the bronchoscope to maintain volume delivery and minimize the development of auto-PEEP [10, 12] are, according to our findings, too optimistic, especially in smaller tubes. For 7.0 or 8.0 mm tubes, for example, a bronchoscope that is 2 mm smaller will result in effective diameters of 4.9 or 5.3 mm, respectively. In both cases, this will lead to significant iPEEP build-up and diminished tidal volume even in a patient with a low respiratory rate and minute ventilation (Fig. 4).

Because the intrinsic PEEP build-up during bronchoscopy under a volume-controlled mode can be dramatic but difficult to estimate without a dedicated end-expiratory occlusion maneuver, we recommend performing bronchoscopy only under a pressure-controlled mode. During pressure-controlled mechanical ventilation, the main detrimental effect of bronchoscopy – a decline in tidal volume – is more easily visible and can be counter-acted by an increase in pressure support, if needed. An additional argument for favoring the added safety of a pressure-controlled mode is that the flow resistance of the endotracheal tube can be considerably higher in a clinical situation, compared to the resistance of unused clean tubes that we have studied here [16]. It is important, however, to reduce the pressure support prior to withdrawing the bronchoscope [12].

Our study demonstrates that the added tube resistance during bronchoscopy can be effectively compensated using full (inspiratory and expiratory) Automatic Tube Compensation (ATC), as opposed to partial ATC as implemented in commercial ventilators [17]. We further show that full ATC can prevent both the reduction in tidal volume and build-up of intrinsic PEEP. However, compensating for bronchoscopy-induced expiratory flow limitation may require lowering airway pressure below atmospheric levels - a function that, to our knowledge, is not available in current commercial ventilators. In addition, accurate compensation requires precise knowledge of the tube resistance. ATC cannot rely on the simplified single-parameter approximation of Eq. 3 but instead requires the full resistance values obtained from Eq. 1 (see Table SI 1). Therefore, these findings provide a compelling rationale to develop ventilators capable of sub-atmospheric expiratory pressure delivery and to incorporate resistance profiles for specific tube– bronchoscope combinations.

## Conclusions

In summary, the effective diameter provides a practical, evidence-based criterion for selecting appropriate combinations of endotracheal tubes and bronchoscopes, rather than relying on oversimplified size recommendations. By knowing the effective diameter, clinicians can anticipate the impact of bronchoscopy on ventilation and adjust ventilator settings accordingly. We provide a freely available predictive framework (https://fabrylab.github.io/Bronchoscopy/) to guide clinicians in selecting ventilator settings that minimize dynamic hyperinflation and hypoventilation, thereby contributing to safer respiratory care during bronchoscopy and improved patient outcomes. Finally, our findings establish a proof-of-principle that automatic tube compensation during bronchoscopy is technically feasible.

## Data Availability

Raw data can be obtained from the corresponding author upon request. The program for predicting ventilation impairments is freely available under MIT license and can be downloaded via the open access repository (https://fabrylab.github.io/Bronchoscopy/)

## Declarations

*Ethics approval and consent to participate:* This study did not involve human participants, data, or identifiable images.

## Consent for publication

Not applicable. This study did not involve human participants, data, or identifiable images.

## Author Contributions

BF, NB, FK, and RF designed the study, CK designed and built the active lung simulator, BF, NB and CK designed and built the ATC ventilator, BF and NB performed the experiments and analyzed the data, JB wrote the software for performing the numerical simulations, BF wrote the first draft of the manuscript, and all authors edited the manuscript.

## Acknowledgements

We thank Saskia Balling for valuable discussions.

---

**Table S1:** Fit parameters of Eq. 1 for describing the pressure drop across the tube for different tube-bronchoscope combinations.

| ETT ∅<br>mm | bronchoscope ∅<br>mm | Deff<br>mm | k1 in,ex<br>mbar | k2 in<br>mbar | k2 ex<br>mbar | rms<br>mbar | MAPE<br>% |
| --- | --- | --- | --- | --- | --- | --- | --- |
| 6.0 | 0.0 | 6.00 | 2.49 | 20.50 | 21.43 | 0.59 | 1.20 |
| 6.5 | 0.0 | 6.50 | 1.59 | 14.53 | 14.75 | 0.42 | 1.30 |
| 7.0 | 0.0 | 7.00 | 1.19 | 12.44 | 10.91 | 0.32 | 1.14 |
| 7.5 | 0.0 | 7.50 | 0.69 | 9.60 | 9.45 | 0.32 | 1.62 |
| 8.0 | 0.0 | 8.00 | 0.47 | 7.32 | 7.30 | 0.27 | 1.89 |
| 8.5 | 0.0 | 8.50 | 0.90 | 5.76 | 5.53 | 0.28 | 1.77 |
| 9.0 | 0.0 | 9.00 | 0.45 | 4.67 | 4.69 | 0.22 | 1.62 |
| 6.0 | 3.8 | 4.64 | 24.39 | 107.17 | 111.22 | 0.50 | 1.91 |
| 6.5 | 3.8 | 5.27 | 10.64 | 56.61 | 58.52 | 0.71 | 1.64 |
| 7.0 | 3.8 | 5.88 | 8.84 | 34.47 | 31.72 | 0.57 | 2.39 |
| 7.5 | 3.8 | 6.47 | 7.12 | 22.82 | 21.32 | 0.75 | 4.30 |
| 8.0 | 3.8 | 7.04 | 3.19 | 16.57 | 15.65 | 0.55 | 3.29 |
| 8.5 | 3.8 | 7.60 | 2.93 | 11.65 | 12.10 | 0.47 | 3.11 |
| 9.0 | 3.8 | 8.16 | 1.11 | 8.86 | 8.91 | 0.30 | 2.52 |
| 6.5 | 5.0 | 4.15 | 56.30 | 206.25 | 225.41 | 0.60 | 2.61 |
| 7.0 | 5.0 | 4.90 | 30.17 | 106.40 | 102.36 | 0.60 | 2.21 |
| 7.5 | 5.0 | 5.59 | 13.65 | 56.64 | 56.67 | 0.69 | 1.29 |
| 8.0 | 5.0 | 6.24 | 8.09 | 30.52 | 29.93 | 0.57 | 1.70 |
| 8.5 | 5.0 | 6.87 | 6.25 | 20.81 | 20.97 | 0.58 | 2.62 |
| 9.0 | 5.0 | 7.48 | 2.86 | 13.36 | 13.56 | 0.46 | 2.03 |
| 7.0 | 5.9 | 3.77 | 109.74 | 464.40 | 475.09 | 0.82 | 1.45 |
| 7.5 | 5.9 | 4.63 | 45.30 | 137.78 | 139.33 | 0.54 | 3.76 |
| 8.0 | 5.9 | 5.40 | 14.49 | 68.87 | 72.43 | 0.75 | 2.20 |
| 8.5 | 5.9 | 6.12 | 12.84 | 36.74 | 35.92 | 0.58 | 3.45 |
| 9.0 | 5.9 | 6.80 | 5.61 | 20.66 | 19.80 | 0.64 | 1.61 |

Deff: effective tube diameter according to Eq. 2
rms: root mean squared error between measured and fitted pressure drop values across the tube (see Fig. 2), excluding all pressure values < 1 mbar
MAPE: mean absolute percentage error between measured and fitted pressure drop values across the tube (see Fig. 2), excluding all pressure values < 1 mbar

**Fig SI 1.**
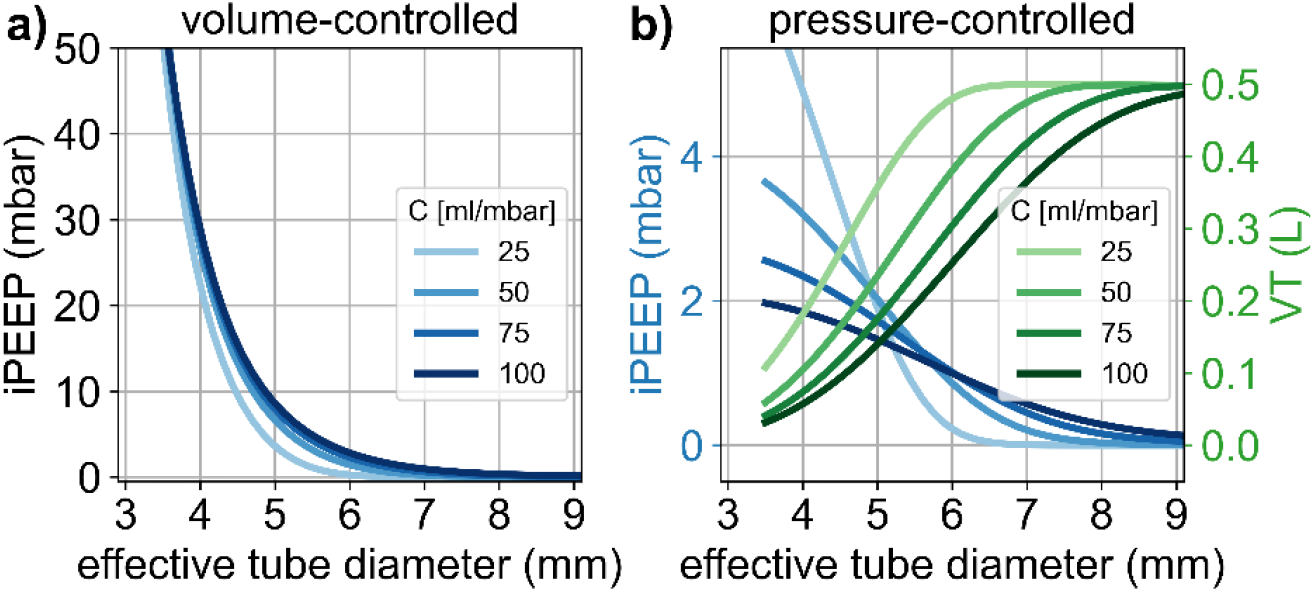
Influence of respiratory system compliance (C = 25, 50, 75 and 100 ml/mbar) on iPEEP and tidal volume. **a)** Intrinsic PEEP build-up during volume-controlled mechanical ventilation, for *V*_T_ = 500 ml, rr = 15/min and t_ex_ = 2.2 s, as a function of the effective tube diameter. **b)** Intrinsic PEEP build-up (blue) and tidal volume changes (green) during pressure-controlled mechanical ventilation for a pressure support (PS) of 10 mbar, RR = 15/min and T_ex_ = 2.2 s, as a function of the effective tube diameter. The PS is adjusted according to PS = 500 ml / *C* so that the maximum tidal volume is 500 ml regardless of compliance. In both graphs, the lines show the results from a numerical simulation based on the resistances of tubes with inserted bronchoscope taken from Eq. 3, for a patient with R_aw_= 2 mbar/(L/s).

**Fig SI 2.**
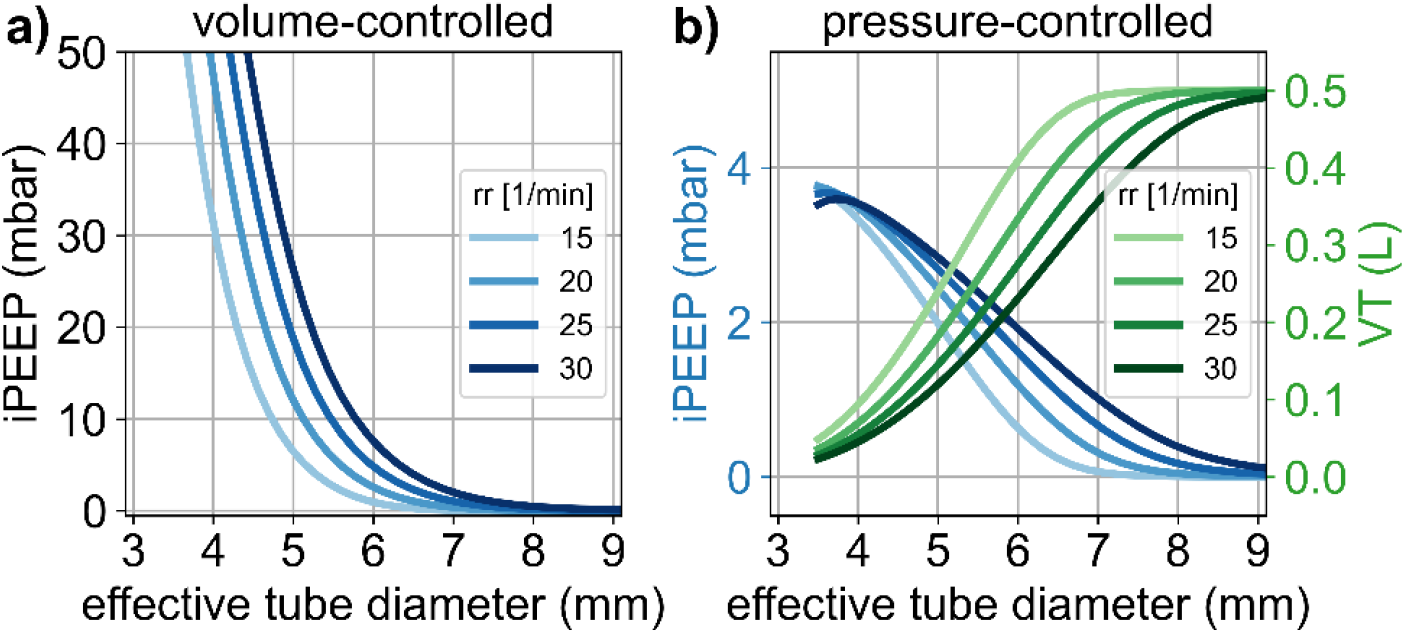
Influence of respiratory rate (RR = 15, 20, 25 and 30 min^−1^) on iPEEP and tidal volume. I:E ratio is set to 1:1.2 in all cases. **a)** Intrinsic PEEP build-up during volume-controlled mechanical ventilation, for *V*_T_ = 500 ml, as a function of the effective tube diameter. **b)** Intrinsic PEEP build-up (blue) and tidal volume changes (green) during pressure-controlled mechanical ventilation for a pressure support of 10 mbar, as a function of the effective tube diameter. In both graphs, the lines show the results from a numerical simulation based on the resistances of tubes with inserted bronchoscope taken from Eq. 3, for a patient with *C* = 50 ml/mbar, R_aw_= 2 mbar/(L/s).

